# Plasma alkaline phosphatase is associated to mortality risk in aortic valve stenosis patients

**DOI:** 10.1101/2022.03.15.22272399

**Authors:** José Pedro L. Nunes

## Abstract

**Background:** aortic valve stenosis is an important clinical condition, with a significant mortality rate in the elderly. Plasma values of alkaline phosphatase (ALP) have been shown to act as a marker of prognosis in different clinical conditions and in the general population.

**Methods:** Plasma levels of alkaline phosphatase were studied in a cohort of patients with aortic valve stenosis, and a five-year survival evaluation was carried out.

**Results:** 24 patients were under study, of which 12 were dead at 5-year follow-up. The median age at baseline evaluation was 79 years (interquartile range, 72-85 years), and 11 patients were of the female sex (13 were male). The median value of ALP, of 83 IU/L, was used to separate patients into two groups; 2 patients died in the group with low ALP values versus 10 patients who died in the group with high ALP values. Using ALP with the same cut-off level, Kaplan Meyer study with log-rank analysis showed a significance level <0.01. Cox regression analysis showed an overall significant result, with a significant level for plasma ALP (significance level 0.03), but not for age, sex or transvalvular gradient (assessed by echocardiography).

**Conclusions:** Elevated plasma ALP is associated to increased mortality risk in aortic valve stenosis patients. This finding may merit evaluation in studies with a larger number of patients.

## Introduction

Aortic valve stenosis is a clinical condition which is growing in importance, since many Human populations across the globe show an increase in their average age and this is a disease that appears mainly in the elderly. The diagnosis of this disease rests mainly, although not exclusively, on echocardiography. Aortic valve stenosis in the elderly is a disease with a significant mortality rate (1). Aortic valve stenosis is currently categorized as severe (in the case of typical high gradient aortic stenosis) if echocardiography shows mean transvalvular gradient >/=40 mmHg, peak velocity >/= 4.0 m/s and valve area </=1 cm^2^ (or </=0.6 cm^2^/m^2^) (2).

Alkaline phosphatase (ALP) is a hydrolase that removes phosphate groups from different types of molecules, acting as an ectoenzyme(being attached to the outer cell membrane). Different isoenzymes are currently recognized: tissue-nonspecific, intestinal, placental, and germ-cell (3). ALP is important for bone mineralization; deficient activity of ALP of genetic origin leads to the clinical condition of hypophosphatasia (4). In a study involving 4155 adults, the authors found an association between ALP and age, waist circumference, body mass index, blood pressure, exercise, alcohol intake, triglycerides, other liver enzymes, cardiovascular disease, arterial hypertension, hypercholesterolemia, and *Diabetes mellitus* (5). Zhong *et al*. found that ALP levels were associated to increased mortality in acute ischemic stroke patients (6). In patients with coronary artery disease, ALP was associated to increased mortality, both in patients with myocardial infarction and *Diabetes mellitus* (7), and in patients who underwent coronary angioplasty with stent implantation (8). Moreover, ALP is currently seen as an independent predictor of mortality in the general population (9).

In the present report, the aim was to study the relation between plasma ALP and survival in a small cohort of aortic valve stenosis patients.

## Methods

Consecutive patients with a diagnosis of aortic valve stenosis were admitted to this study, carried out in a general Cardiology outpatient clinic. Baseline evaluation included echocardiography, electrocardiography and blood tests, including ALP.

In the present report, 5–year (60 months) survival was under study, and this was established prospectively by the study of electronic health records, after a minimum period of 55 months for each patient had passed from the initial evaluation. In the case of dead patients, the date of death was recorded, when available, or alternatively the date of the last observation of each patient was used. In the case of patients not known to be dead, censoring was carried out in the date of the last observation. No attempt was made to study the causes of death.

The two groups of patients (dead or alive) were compared by means of (the non-parametric) Mann-Whitney U test, considering age, mean left ventricular/aortic gradient (as measured by echocardiography; mean transvalvular gradient) and plasma ALP.

The median value for ALP in the present cohort under study (83 International Units/ Liter - IU/L) was used as cut-off, and Kaplan-Meyer study was carried out. The comparison between groups was made using the log-rank test.

Cox-proportional hazards survival modelling was used. Covariates included gender, age, plasma ALP, and mean transvalvular gradient. The relative risk for mortality was calculated by dividing the cases in two groups according to the median ALP value.

A significance level of 0.05 or lower was considered statistically significant. Data analysis was performed using the SPSS 26 software program, from IBM (Amonk, NY, USA), except for relative risk calculation, for which the MedCalc online calculator was used (available at https://www.medcalc.org/calc/relative_risk.php).

The present research project was approved by the ethics committee of our institution, and individual informed consent was obtained from every patient admitted to the study.

## Results

Two patients were not enrolled in the study due to lack of willingness to sign the informed consent form. A total number of 24 patients were under study, out of an initial number of 27 patients. Three patients were excluded due to not having performed blood studies at our institution. Baseline data are presented in Table 1.

**Table 1.** Baseline characteristics of the aortic valve stenosis patients cohort under study. ALP – alkaline phosphatase. IU/L – International Units/ Liter.

|  | Median | Interquartile range |
| --- | --- | --- |
| Age (years) | 79 | 72-85 |
| Transvalvular gradient<br>(mm Hg) | 35 | 31-46 |
| Plasma ALP (IU/L) | 83 | 57-115 |
| Sex | 11 Female patients | 13 Male patients |

A total number of 7 patients underwent aortic valve intervention during the 5 year follow up time (surgery in 6 cases and percutaneous valve implantation in the remaining case). These 7 patients had, cumulatively: a severe degree of valve stenosis (as evaluated by echocardiography); willingness to allow valve intervention; acceptance from the part of the surgical/ cardiac hemodynamic team. These seven patients were alive at end follow-up.

Concerning electrocardiography, two patients had atrial fibrillation, one had atrial flutter and the remaining were in sinus rhythm.

At 5-year follow-up, 12 patients were dead and 12 were alive (one patient in this latter group had a 56-months follow-up, unlike all others, with a 60-month follow-up). Two patients died in the group with low ALP values, versus 10 patients who died in the group with high ALP values

Mann-Whitney test showed that age (p<0.01) and plasma ALP (p<0.01) were significantly higher in dead patients, when compared to patients living at 5-year follow-up time; the same did not happen in the case of mean transvalvular gradient (p=0.13).

As can be seen in Figure 1, patients with an ALP plasma level above the median value had a marked mortality rate, unlike the other patients. Kaplan-Meyer study with log-rank test showed an overall significance level <0.01.

**Figure 1.**
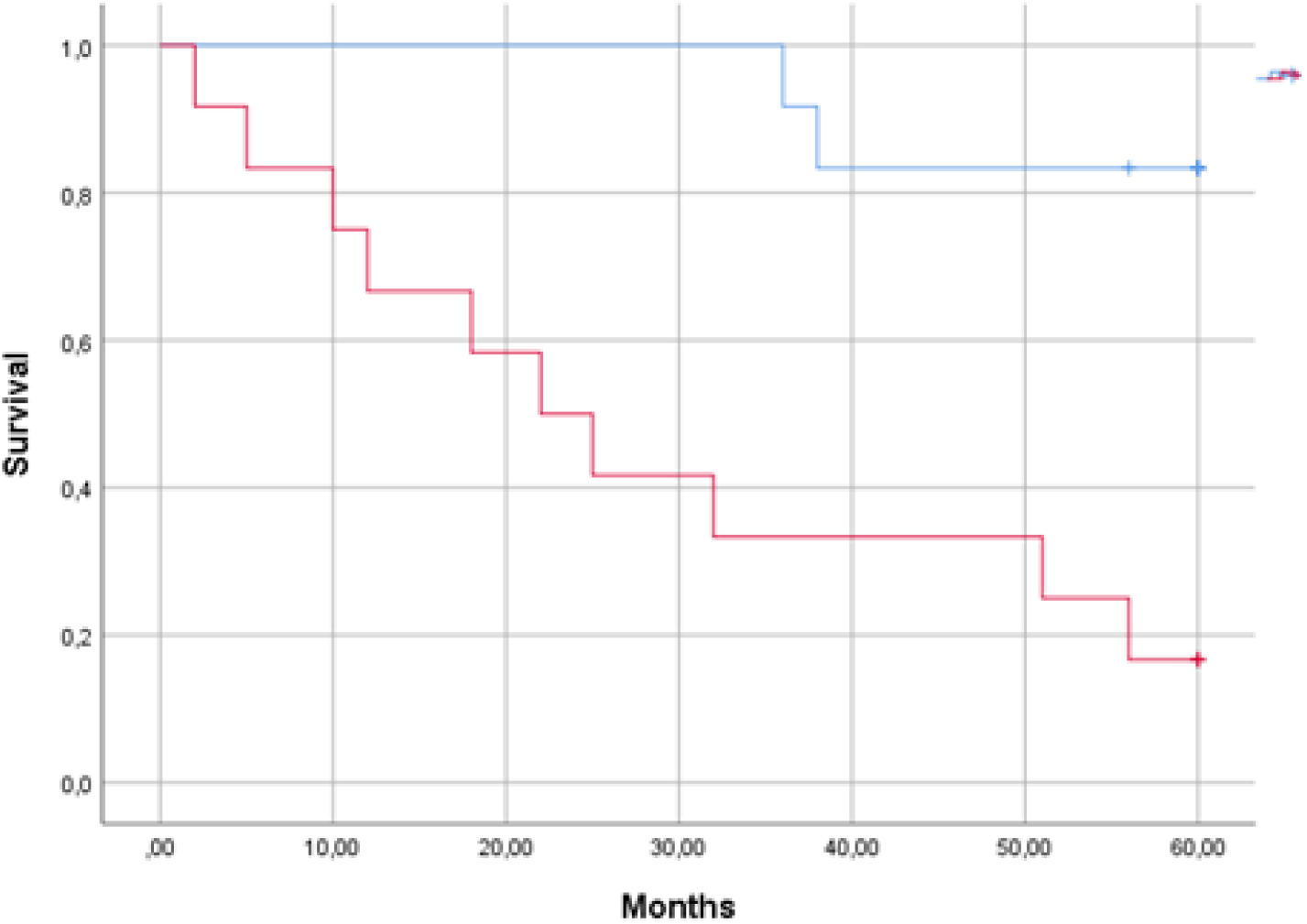
Kaplan-Meyer survival curves for aortic valve stenosis patients with plasma alkaline phosphatase below (top line, 12 patients) or above (bottom line, 12 patients) the median value of 83 IU/L.

A relative risk of 5.0 was seen for mortality for patients with increased ALP levels, when compared to patients with lower levels (95% confidence interval of 1.4-18.2).

Cox regression analysis showed an overall significant result, with a significant level for plasma ALP (Table 2). Similar findings were seen after inserting an additional variable (valve intervention) in the model (the new variable was not significant in this model; data not shown).

**Table 2.** Significance levels for Cox regression analysis of the aortic valve stenosis patients cohort under study. ALP – alkaline phosphatase. IU/L – International Units/ Liter.

|  |  |
| --- | --- |
| Overall | <0.01 |
| Sex | 0.47 |
| Age (years) | 0.17 |
| Trans-valvular gradient<br>(mm Hg) | 0.26 |
| Plasma ALP (IU/L) | 0.03 |

## Discussion

In the present report, elevated plasma ALP was shown to be able to identify aortic valve stenosis patients with a high mortality rate. Due to the wide availability of this biomarker, ALP could be established as a useful aid in the management of this type of patients. As stated above, elevated ALP has been shown to act as a negative prognostic factor both in patients with heart disease and in the general population (7, 8, 10), and ALP is currently seen as an independent predictor of mortality in the general population (9).

The mechanism underlying the increased mortality seen in patients with elevated ALP levels is unclear at the present stage. Vascular calcification is frequently presented as a consequence of increased ALP activity, since the enzyme hydrolyzes pyrophosphate, an inhibitor of vascular calcification (10). In the case of aortic valve stenosis patients, increased valve calcification could exist in the setting of increased ALP levels. Clark-Greuel *et al*. showed that ALP gene expression was increased in some but not all cases of Human calcified valves (11). These authors also showed that ALP was increased in experimental (cell-culture) stimulation of calcification of aortic valve interstitial cells.

ALP generates inorganic phosphate (9); ALP is present in myocardial capillary endothelial cells (12); ALP may modulate inflammation (9), which may also play a part in the context under study. The possibility that changes in the liver associated to hemodynamic changes lead to increased ALP levels cannot be ruled out at the present stage. ALP levels could vary in the setting of both liver and systemic diseases.

Several types of biomarkers have been studied in aortic valve stenosis patients, including cardiac troponins and natriuretic peptides, and have been shown to correlate with patient mortality (13). A major interest of biomarker studies in aortic valve stenosis, apart from evaluating prognosis, would be to guide the ideal time frame for valve intervention, a topic possibly to be explored in clinical trials, in the case of ALP.

Furthermore, one may speculate that the study of the intrinsic mechanisms by which increased ALP is associated to increased mortality could allow the finding of useful interventions to decrease mortality in these patients, as well as in others.

## Study limitations

The present study has significant limitations, the main of which is that the small dimension of the sample limits the strength of conclusions. This small cohort includes only patients with a relatively advanced age (range of 65-93 years at baseline evaluation), and therefore the findings may not be applicable to younger patients.

## Conclusions

Elevated plasma ALP is associated to increased mortality risk in aortic valve stenosis patients. This finding may merit evaluation in studies with a larger number of patients.

## Data Availability

All data produced in the present study are available upon reasonable request to the author

